# Limited Educational Resources and Infrequent Contact Hinders Dietitian-Led Weight Management Success

**DOI:** 10.64898/2026.03.11.26348045

**Authors:** Ashleigh Oliveira, Manabu T Nakamura

## Abstract

Due to a high prevalence of obesity-related chronic diseases and an increasing use of injectable weight loss medications, a need for dietary consultations by registered dietitian nutritionists (RDNs) has been increasing. However, there is a paucity in RDNs who provide weight management services. The objective of this study is to gather insight on the dietary interventions and barriers in weight loss practice as reported by RDNs specializing in weight management. The survey contained 33 questions and four domains (current practice diet interventions and protocols, client demographics, practitioner demographics, and barriers). Descriptive statistics were used to evaluate results. Survey participants, RDNs (N=739), were recruited from Weight Management Dietetic Practice Group of the Academy of Nutrition and Dietetics. Weight management occupies most (69%) of surveyed RDNs’ work time. App use was the most common method of food record collection (n=299), and food record accuracy was the most prevalent cited barrier to weight loss magnitude (n=233). Most (n=253) RDNs reported seeing their typical client once a month with an average continued engagement of one month being the most prevalent (n=197). Lastly, most RDNs (81%, n=595) do not have a customary program or diet they refer clients to. In conclusion, improvements in technologically enhanced platforms for accurate diet record collection and patient education may benefit RDNs weight management practice and further research into reasons for patient attrition from weight loss counselling is warranted.

## 1. Introduction

The prevalence of obesity in the United States (US) increased from 30.5% in 2010 to 40.3% in 2023 [1, 2]. Excess weight is associated with comorbidities that can be deleterious to quality of life, but these can be ameliorated with weight loss [3]. In the US, Registered Dietitian Nutritionists (RDNs) are the primary interventionists assigned to patients who are seeking weight loss before more invasive measures are considered; however, the supply of nutrition professionals falls short of meeting the demand, with only 112,000 actively credentialed RDNs available to serve over 100 million adults living with obesity [4, 5].

Dietitian intervention leads to better weight loss outcomes than usual or no care [6]. Potential reasons for better success were indicated in a systematic review of 62 randomized controlled trials that demonstrated a greater effect size in interventions that had more touch points with the RDN and those with a longer treatment period [6]. Most of the nutrition interventions in this systematic review can be categorized as cognitive behavioral therapy, health behavior change support system, alternate day fasting, or calorie restriction, but no specific diet intervention can be quantified as more or less effective [6].

In a survey of 100 RDNs, certified clinical nutritionists, and certified nutrition specialists, participants were asked about solutions that would be the most helpful to facilitate their patients’ weight loss [7]. Most indicated greater or added reimbursement from insurance companies would aid in the treatment of their patients (71/100) [7]. Other popular solutions included training for the entire patient care team on obesity care (50/100), compensation for weight-loss related training of providers (36/100), and having an online weight loss program to refer patients to (22/100) [7].

Innovations in technology have changed the way that patients interact with health care providers and receive their health-related information. Telehealth is a promising solution for patients located remotely and for those with limited time and resources to access a healthcare facility [8, 9]. Additionally, the internet and smart phone applications are becoming more accessible as a resource for health-related information, diet, and weight tracking. However, some studies indicate the efficacy of interventions delivered exclusively by telehealth is lesser than those delivered in-person and there are insurance coverage limitations and concerns over quality and reliability [6, 10].

Notably in recent years, weight loss medications such as glucagon-like peptide-1 (GLP-1) receptor agonists are increasing in demand, but this does not make the role of nutrition counselling obsolete. GLP-1 receptor agonists demonstrate significant weight loss in most patients; however, many patients choose to stop taking the medication due to cost or side effects and experience a regain of lost weight [11]. Additionally, the high expense of these weight loss medications and the potential long-term reliance have encouraged insurance companies to require a lifestyle and diet intervention in exchange for medication coverage [11–13]. Dietitians with expertise in weight loss fill a pivotal role by providing the lifestyle and diet interventions to support sustainable long-term results.

In summary, the demand for diet support for weight management is increasing, most notably for RDNs who can help patients make long-term diet and lifestyle changes. The way we access healthcare is changing due to technology enhancements and health insurance policy changes. While research indicates more contact and longer interventions to be more successful, there is a deficit in information regarding the specific diets, programs, and technologies used by RDNs with weight loss patients. Furthermore, due to recent changes in weight loss medications and insurance coverage, RDNs’ roles in these arenas requires elucidation. Therefore, the objective of this study was to evaluate the dietary interventions and tools currently used by RDNs for weight management, including their perceived effectiveness and the challenges they face in achieving client weight loss success.

## 2. Methods and Materials

### 2.1 Survey Development

A draft survey was developed based on two studies with similar research objectives. The majority of survey questions were modeled from the Clinician Apps Survey (CAS) which was developed to assess how clinicians who treat patients with obesity and/or diabetes use apps in their practice [14]. The CAS survey contains 37 questions of which the included domains are client and clinician demographics, technology use, client assessments, counseling strategies, technology use with clients, and factors affecting technology use by both the clinician and client. Protocol questions regarding the number of appointments, follow up procedures, and details of the specific weight management programs were adapted from a survey created to identify management protocols used by RDNs for clients with obesity in the British Dietetic Association [15].

The drafted survey contained 37 questions which were then reviewed by an expert panel for sufficiency in addressing the primary objective, appropriate language, and clarity. The panel consisted of thirteen participants including nutrition graduate student researchers (n=2), RDNs (n=4), students of the didactic program of dietetics (n=4), nutrition educators (n=2), and a representative member (n=1) of the Weight Management Dietetic Practice Group (WMDPG).

The resulting survey contained 33 questions in total including 18 on current diet interventions and protocols, nine on client demographics and characteristics, and six on clinician demographics. Current practice diet interventions included methods and frequency of diet assessment, weight monitoring, use of technology, and weight management programs and apps. In addition, barriers to client and practitioners are inquired within each category. The survey was production tested by the panel prior to being placed into production mode.

### 2.2 Survey Administration

The study was deemed exempt by the University of Illinois Institutional Review Board under federal regulation 45 46.101 (b) Code of Federal Regulations (IRB Protocol ID 24173). A consent cover letter was provided to all participants. RDNs were recruited from the WMDPG. Members of the practice group are RDNs, dietetic technicians, dietetic students, or eligible candidates of the Registration Examination for Dietitians or Dietetic Technicians who have opted to join this professional interest group. Exclusion criteria included being younger than 18 years old, having less than 1 year of weight loss practice as an RDN, and not having a currently active RDN credential as provided by the Commission. Potential participants were provided with a brief survey description and a Research Electronic Data Capture link disseminated by email blast, the monthly newsletter, and a posting on the discussion board. Approximately, 3,500 members opt to receive emails from the WMDPG. The survey link directed participants to an electronic consent required for participation. The first survey question reiterated confirmation of the inclusion criteria that was required to be answered to move to the next question. The final question of the survey provided an optional submission to a random draw for one of two $100 Amazon electronic gift cards. From date of release, respondents had four weeks to complete the survey. Participants could revisit questions while within the survey and save answers if they desired to return later. Only one submission was possible per provided user email. All identifiers were removed from analyzed survey files by the primary researcher.

### 2.3 Statistical Analyses

Descriptive statistical analyses were conducted using SPSS 26.0 (IBM Corp. Released in 2019. IBM SPSS Statistics for Windows, Version 24.0. Armonk, NY, USA: IBM Corp.)

## 3. Results

### 3.1 Response

The survey was administered and left open for four weeks, 767 complete responses were collected, 739 responses were kept for analysis. Twenty-eight responses were removed due to incompatible education with reported credentials (RDN with highest testified education as high school or some college). Survey participants that did not self-report as being a currently credentialed RDN were excluded from the study using skip logic terminating the survey. All responses were self-reported. All respondents indicated that they allocated a certain percentage of time to weight management practice, with an average of 69% (n=510).

### 3.2 Dietitian Demographics and Practice Settings

Demographic information of RDN respondents can be found in Table 1. Most survey respondents were white (89%, n=660), female (84%, n=618), 26-35 years old (342, n=46%) with a master’s degree (70%, n=520). Most participants’ reported years of experience as a RDN was 5-10 years (69%, n=510). Current practice settings were divided and some reported working among multiple arenas including, but not limited to outpatient (31%, n=231), private practice (24%, n=180), food service (22%, n=164), inpatient (22%, n=162), and nutrition education (22%, n=161).

**Table 1.** Demographics and Practice Setting for Participant Dietitians of the Weight Management Dietetic Practice Group (N=739)

| **Question** | **Results** | **n** | **%** |
| --- | --- | --- | --- |
| **Gender** | Female | 618 | 83.6% |
|  | Male | 106 | 14.3% |
|  | Prefer not to answer | 15 | 2.0% |
| **Age, years** | 18-25 | 48 | 6.5% |
|  | 26-35 | 342 | 46.3% |
|  | 36-45 | 304 | 41.1% |
|  | 46-55 | 29 | 3.9% |
|  | 56-65 | 13 | 1.8% |
|  | 66-75 | 3 | 0.4% |
| **Race** | White/Caucasian | 660 | 89.3% |
|  | Black/African American | 37 | 5.0% |
|  | Hispanic or Latino | 16 | 2.2% |
|  | Asian/Pacific Islander | 12 | 1.6% |
|  | I prefer not to answer | 9 | 1.2% |
|  | Native American | 5 | 0.7% |
| **Highest level of education** | Master's degree | 520 | 70.4% |
|  | 4-year college degree | 128 | 17.3% |
|  | Doctoral degree | 65 | 8.8% |
|  | Judicial/Medical | 26 | 3.5% |
| **Professional credentials and advanced training** | Registered Dietitian | 739 | 100.0% |
|  | Nutritionist | 108 | 14.6% |
|  | Registered Nurse or Licensed Practicing Nurse | 76 | 10.3% |
|  | Nurse Practitioner | 48 | 6.5% |
|  | Physician's Assistant | 36 | 4.9% |
|  | Medical Doctor | 6 | 0.8% |
|  | Others, not listed: |  |  |
|  | Certified Diabetes Care and Education Specialist | 5 | 0.7% |
|  | Exercise Physiologist | 4 | 0.5% |
|  | Licensed Acupuncturist | 1 | 0.1% |
| **Years practicing as a dietitian** | < 1 year | 20 | 2.7% |
|  | 1-5 years | 147 | 19.9% |
|  | 5-10 years | 510 | 69.0% |
|  | >10 years | 62 | 8.4% |
| **Practice setting** | Outpatient or ambulatory care | 231 | 31.3% |
|  | Private practice, outpatient | 180 | 24.4% |
|  | Diet planning in food service | 164 | 22.2% |
|  | Inpatient | 162 | 21.9% |
|  | Nutrition education | 161 | 21.8% |
|  | Others, not listed: |  |  |
|  | Industry | 16 | 2.2% |
|  | Research | 3 | 0.4% |
| **Disease types treated** | Weight loss | 739 | 100.0% |
|  | Pre and post bariatric care | 247 | 33.4% |
|  | Cardiovascular Disease | 214 | 29.0% |
|  | Hypertension | 204 | 27.6% |
|  | Gestational Diabetes | 200 | 27.1% |
|  | Chronic Kidney Disease/Dialysis patients | 188 | 25.4% |
|  | Prediabetes and Type 2 Diabetes | 178 | 24.1% |
|  | Type 1 Diabetes Mellitus | 176 | 23.8% |
|  | In-patient care | 162 | 21.9% |
|  | Enteral and parenteral nutrition | 158 | 21.4% |
| **Participant client age** | < 10 years old | 242 | 32.7% |
|  | 11-18 years old | 290 | 39.2% |
|  | 19-29 years old | 406 | 54.9% |
|  | 30-49 years old | 399 | 54.0% |
|  | 50-69 years old | 368 | 49.8% |
|  | 70-84 years old | 76 | 10.3% |
|  | 85+ years old | 35 | 4.7% |

Responses to demographic questions provided by dietitians of the Weight Management Dietetic Practice Group (WMDPG) of the Academy of Nutrition and Dietetics. The total (n) number of dietitians who selected the answer and the percentage (%) of dietitians out of the total (N=739).

RDNs reported treating clients of in multiple age brackets majority treating ages 19-29 years old (55%, n=406) and 30-49 years old (54%, n=399). On opposite ends of the age spectrum, 33% (n=242) of RDNs reported treating children of less than 10 years as opposed to 15% (n=111) treating patients seventy and older years of age. All practitioners reported treating patients for weight loss (100%, N=739), while other conditions associated to weight such as pre and post bariatric care (33%, n=247), cardiovascular disease (29%, n=214), hypertension (28%, n=204) and pre- and type 2 diabetes (24%, n=178) were relatively split.

### 3.3 Methods of Diet Assessment

Methods of diet assessment by RDN respondents can be found in Table 2. Almost all respondents claimed to routinely assess patient diet (96%, n=710). For those that did not assess diet (4%, n=29), the reported reasons were feeling that clients did not report accurate diet information (n=10), clients did not return diet records (n=9), RDNs did not think diet assessment was useful (n=7), and RDNs did not have enough time (n=3). RDNs reported using multiple means of recording diet intake including self-reported food diaries (n=295), interviews of usual intake (n=291), 24-hour recalls (n=209), and food frequency questionnaires (n=169). Forty-six RDNs voluntarily commented that they use photo diaries. The most common frequency of diet assessment collection was once a month (29.6%, n=219), followed closely by once a week (26%, n=189), with few claiming once a year or less (4%, n=26). RDNs recommended a variety of tracking tools to clients for diet assessment, but were majority smartphone apps (41%, n=299), computer-based processers (36%, n=263), online programs/websites (32%, n=236), and some recommending pen and paper (26%, n=189).

**Table 2.** Methods of Diet Assessment for Participant Dietitians of the Weight Management Dietetic Practice Group (N=739)

| **Question** | **Results** | **n** | **%** |
| --- | --- | --- | --- |
| **Routinely assess diet** | Yes | 710 | 96.1% |
|  | No | 29 | 3.9% |
| **Reasons for not routinely assessing diet (n=29)** | Information received from clients is unreliable. | 10 | 34.5% |
|  | Clients do not return diet records when requested. | 9 | 31.0% |
|  | I don't think it is useful to assess diet intake. | 7 | 24.1% |
|  | I don't have enough time. | 3 | 10.3% |
| **Methods of diet assessment** | Self-reported Food Diary | 295 | 39.9% |
|  | Interview of Usual Diet Intake | 291 | 39.4% |
|  | 24-hour Food Recall | 209 | 28.3% |
|  | Food Frequency Questionnaire | 169 | 22.9% |
|  | Others, not listed: |  |  |
|  | Photos of meals | 46 | 6.2% |
| **Frequency of diet assessment** | Once a month | 219 | 29.6% |
|  | Once a week | 189 | 25.6% |
|  | Once every 3 months | 156 | 21.1% |
|  | Once every 6 months | 149 | 20.2% |
|  | Once a year | 13 | 1.8% |
|  | < 1 time per year | 7 | 0.9% |
|  | I never collect dietary intake | 6 | 0.8% |
| **Methods of diet assessment collection** | Smartphone app | 299 | 40.5% |
|  | Computer-based Word Processing (i.e., Word, Excel) | 263 | 35.6% |
|  | Online program or website | 236 | 31.9% |
|  | Pen and paper | 189 | 25.6% |
|  | None, I don't recommend tracking tools for clients. | 6 | 0.8% |

Responses to diet assessment methods questions provided by dietitians of the Weight Management Dietetic Practice Group (WMDPG) of the Academy of Nutrition and Dietetics. The total (n) number of dietitians who selected the answer and the percentage (%) of dietitians out of the total (N=739).

### 3.4 Methods of Weight Assessment

Methods of weight assessment by RDN respondents can be found in Table 3. Similar to results of the methods of diet assessment, 96% (n=710) of RDNs reported routinely assessing weight with only 29 (4%) respondents claiming they did not, however these RDNs were not largely overlapping with only two respondents claiming they do not routinely assess diet and weight. The reasons for not routinely assessing weight included feeling that clients did not want to be weighed (41%, n=12), feeling it is not useful to assess weight (41%, n=12), and not having enough time to assess weight (17%, n=2). Most RDNs ask clients to track their own weight (96%, n=706). For those that did not, RDNs reported negative effects of self-weighing on client mood (n=16), clients do not want to weight themselves (n=6), they don’t think this is a useful practice (n=6), lack of reliable home scales (n=5), and that clients do not have time (n=2). A few voluntarily elaborated on this question expressing concern of enforcing disordered thoughts and habits (n=3), and that their clients had limited mobility to weight themselves (n=1).

**Table 3.** Methods of Weight Assessment for Participant Dietitians of the Weight Management Dietetic Practice Group (N=739)

| **Question** | **Results** | **n** | **%** |
| --- | --- | --- | --- |
| **Routinely assess weight** | Yes | 710 | 96.1% |
|  | No | 29 | 3.9% |
| **Reasons for not routinely assessing weight (n=29)** | Clients do not want to be weighed. | 12 | 41.4% |
|  | I don't think it is useful to assess weight. | 12 | 41.4% |
|  | I don't have enough time. | 5 | 17.2% |
| **Clients asked track their own weight** | Yes | 706 | 95.5% |
|  | No | 33 | 4.5% |
| **Reasons clients not asked to track their own weight (n=33)** | Weighing has negative effects on client mood. | 16 | 48.5% |
|  | Clients do not want to weight themselves. | 6 | 18.2% |
|  | I don't think it is useful to self-monitor weight. | 6 | 18.2% |
|  | Home scales are not reliable. | 5 | 15.2% |
|  | Clients do not have time. | 2 | 6.1% |
|  | Others, not listed: |  |  |
|  | Enforces disordered thoughts and habits | 3 | 0.4% |
|  | Limited mobility | 1 | 0.1% |

Responses to weight assessment methods questions provided by dietitians of the Weight Management Dietetic Practice Group (WMDPG) of the Academy of Nutrition and Dietetics. The total (n) number of dietitians who selected the answer and the percentage (%) of dietitians out of the total (N=739).

### 3.5 App and Technology Use

The reported use of apps and technology of RDNs’ clients can be found in Table 4.

**Table 4.**
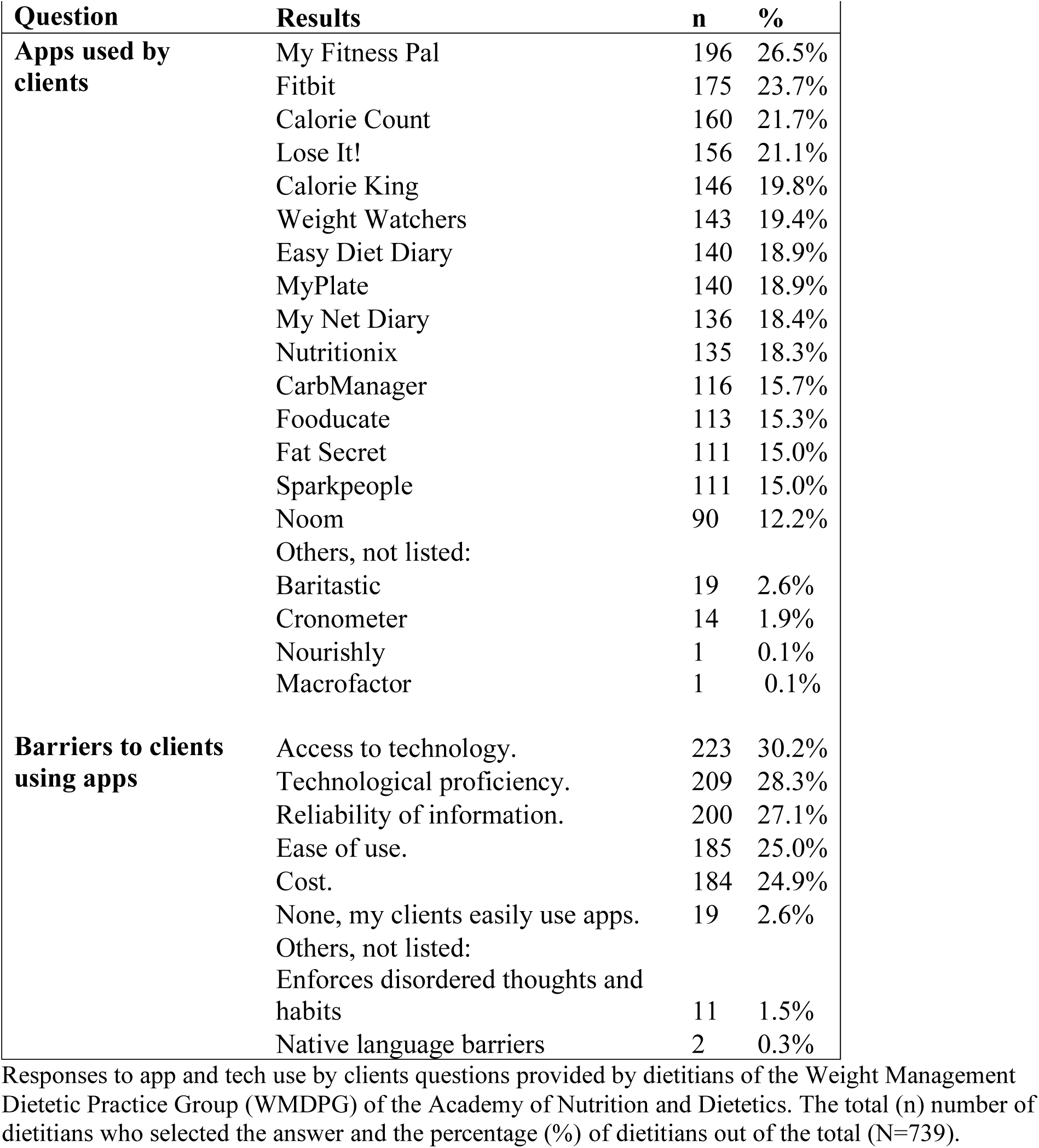
Apps and Technology Use by Clients for Participant Dietitians of the Weight Management Dietetic Practice Group (N=739)

Several apps were reportedly used by the RDNs’ clients with the majority being My Fitness Pal (27%, n=196), Fitbit (24%, n=175), Calorie Count (22%, n=160), Calorie King (20%, n=146), and Weight Watchers (19%, n=143). When enquired about barriers their clients face in using apps, most reported limitations in access to technology (30.2%, n=223), technological proficiency (28%, n=209), unreliable information provided by the application (27%, n=200), ease of use (25%, n=185), and cost (25%, n=184). Only 19 (3%) RDNs reported that there were no barriers to their clients using apps for weight loss. Other barriers to using apps voluntarily elaborated on included enforcing disordered thoughts and habits (2%, n=11) and a lack of apps provided in their clients’ native language (0.3%, n=2).

The reported use of apps and technology used by RDNs themselves can be found in Table 5. All RDNs reported using some kind of digital device for work and many used multiple, majority using a laptop or desktop (86%, n=634), tablet or iPad (75%, n=553), and/or smartphone (75%, n=551). Almost all RDNs use websites and/or apps for work (95%, n=699) and many use multiple means of health-related media at work including but not limited to smartphone apps (33%, n=243), websites (30%, n=221), tablet or iPad apps (27%, n=198), YouTube (26%, n=190), and Facebook (25%, n=185).

**Table 5.** Apps and Technology Use by Dietitians for Participant Dietitians of the Weight Management Dietetic Practice Group (N=739)

| **Question** | **Results** | **n** | **%** |
| --- | --- | --- | --- |
| **Digital devices used for work** | Laptop or desktop computer | 634 | 85.8% |
|  | Tablet or iPad | 553 | 74.8% |
|  | Smartphone | 551 | 74.6% |
| **Apps and websites used for work** | Yes | 699 | 94.6% |
|  | No | 40 | 5.4% |
| **Health-related media used for work** | Smartphone apps | 243 | 32.9% |
|  | Online programs/websites | 221 | 29.9% |
|  | Tablet or iPad apps | 198 | 26.8% |
|  | YouTube Videos | 190 | 25.7% |
|  | Facebook | 185 | 25.0% |
|  | Educational materials, fliers, brochures | 163 | 22.1% |
|  | Videos (DVDs, CDs) | 158 | 21.4% |
|  | Twitter | 149 | 20.2% |
|  | PowerPoint | 144 | 19.5% |
|  | Instagram | 131 | 17.7% |
|  | Pinterest | 129 | 17.5% |

Responses to app and tech use by dietitians questions provided by dietitians of the Weight Management Dietetic Practice Group (WMDPG) of the Academy of Nutrition and Dietetics. The total (n) number of dietitians who selected the answer and the percentage (%) of dietitians out of the total (N=739).

### 3.6 Weight Management Outcomes and Protocols

Details of the RDNs’ weight management outcomes are in Table 6. RDNs reported spending an average of 69% of their work on weight management. Four participants reported spending less than 10%, unanimously reporting weight management protocols to be ineffective. Most RDNs reported that they did not have a standard weight management program for their clients (81%, n=595). Of those who did rely on a specific program (19%, n= 144), most claimed it was a low-carbohydrate diet (n=57), the Obesity Medicine Program diet (n=36), or a Mediterranean diet (n=17) [16]. The average number of consultations provided by RDNs to clients seeking weight loss was mostly once a month (34%, n=253), once a week (25%, n=183), or once every 3 months (21%, n=154). The average length of continued treatment of a client seeking weight loss was mostly one month (26%), with a fairly even split among three months (22%, n=163), six months (22%, n=161), and 1 week (21%, n=153). Some RDNs optionally reported barriers to clients’ weight loss magnitude included clients having limited income (n=65), undiagnosed eating disorders (n=36), food insecurity (n=26), depression (n=23), insurance coverage (n=19), and lack of adherence to prescribed diet (n=16).

**Table 6.**
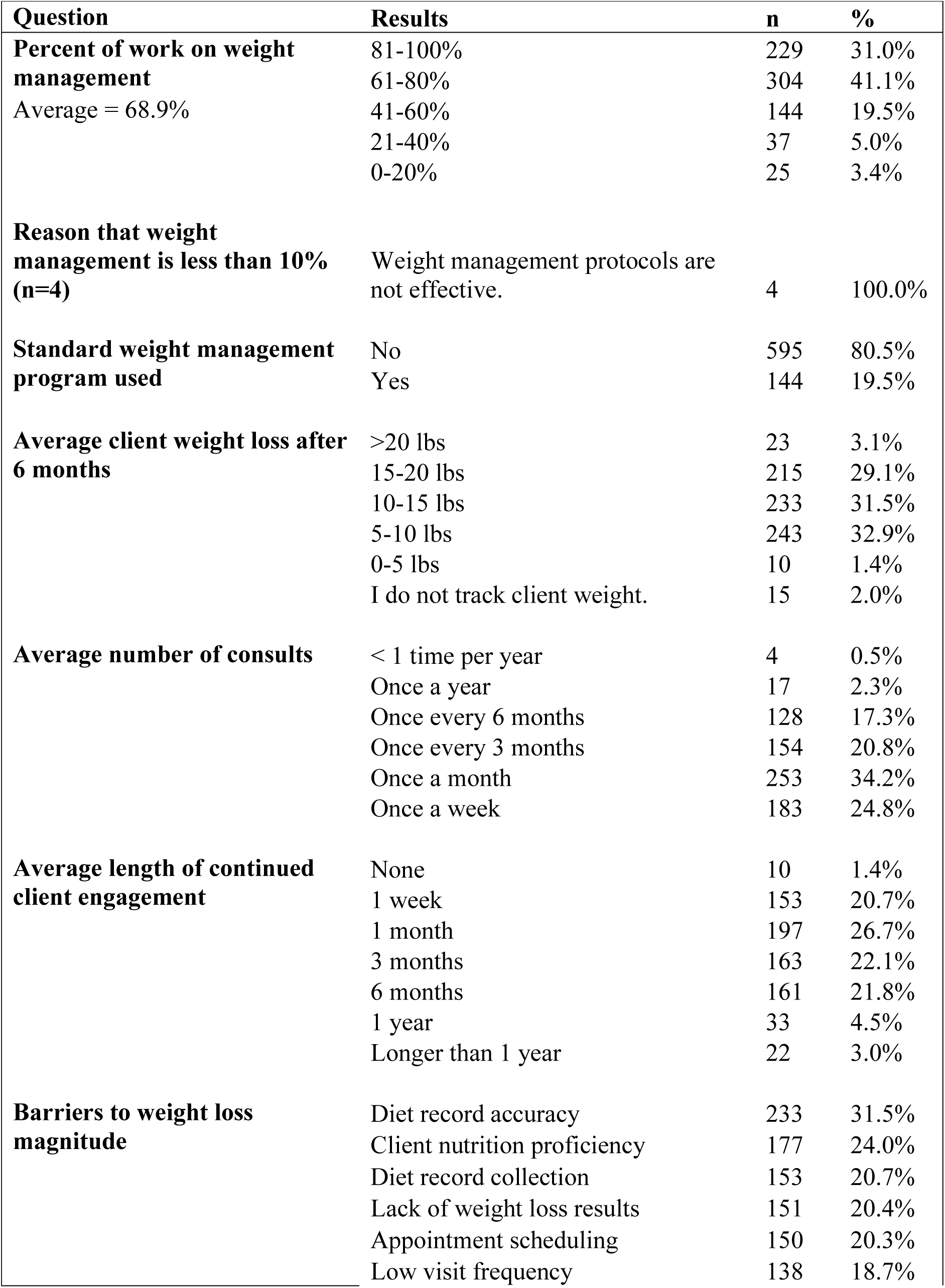
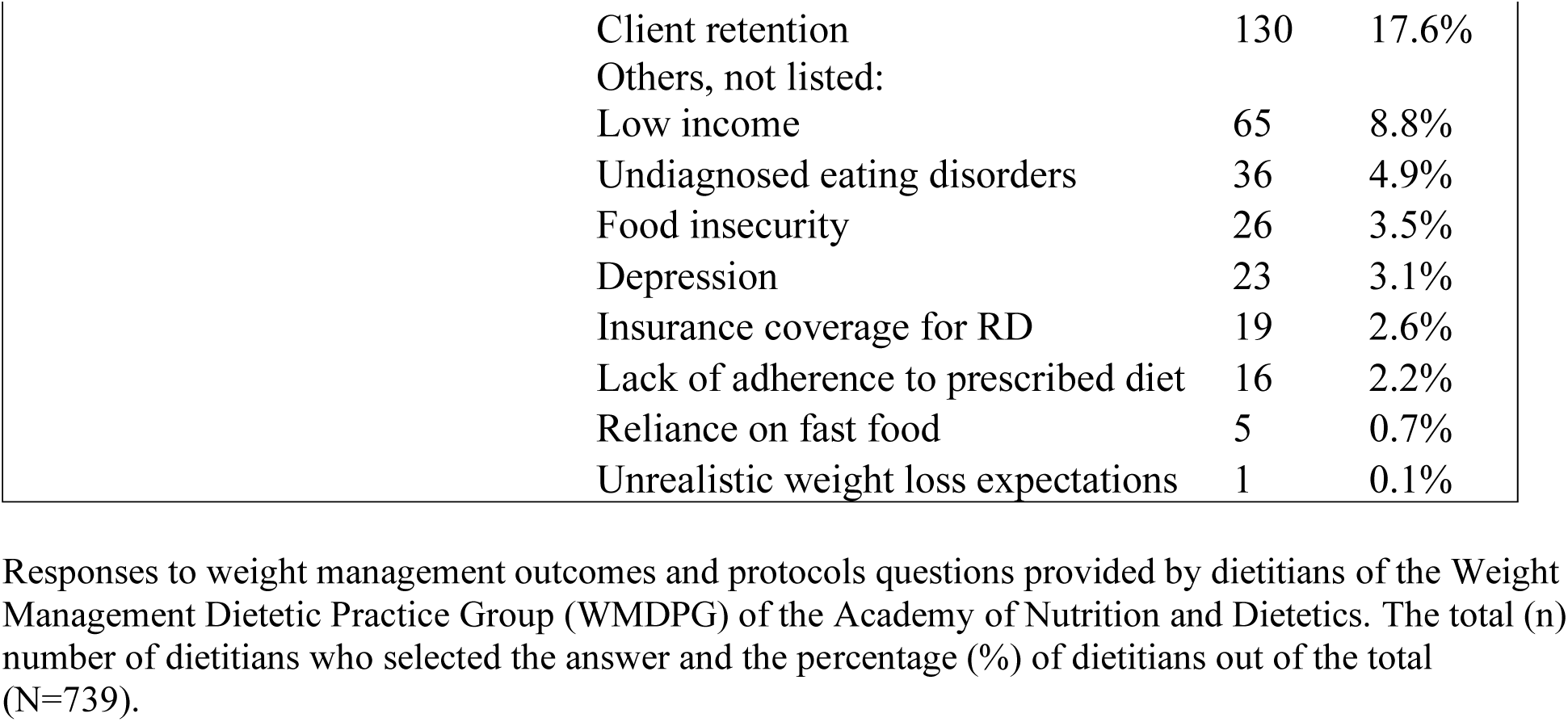
Weight Management Outcomes and Protocols for Participant Dietitians of the Weight Management Dietetic Practice Group (N=739)

| **Question** | **Results** | **n** | **%** |
| --- | --- | --- | --- |
| **Percent of work on weight management**<br>Average = 68.9% | 81-100% | 229 | 31.0% |
|  | 61-80% | 304 | 41.1% |
|  | 41-60% | 144 | 19.5% |
|  | 21-40% | 37 | 5.0% |
|  | 0-20% | 25 | 3.4% |
| **Reason that weight management is less than 10% (n=4)** | Weight management protocols are not effective. | 4 | 100.0% |
| **Standard weight management program used** | No | 595 | 80.5% |
|  | Yes | 144 | 19.5% |
| **Average client weight loss after 6 months** | >20 lbs | 23 | 3.1% |
|  | 15-20 lbs | 215 | 29.1% |
|  | 10-15 lbs | 233 | 31.5% |
|  | 5-10 lbs | 243 | 32.9% |
|  | 0-5 lbs | 10 | 1.4% |
|  | I do not track client weight. | 15 | 2.0% |
| **Average number of consults** | < 1 time per year | 4 | 0.5% |
|  | Once a year | 17 | 2.3% |
|  | Once every 6 months | 128 | 17.3% |
|  | Once every 3 months | 154 | 20.8% |
|  | Once a month | 253 | 34.2% |
|  | Once a week | 183 | 24.8% |
| **Average length of continued client engagement** | None | 10 | 1.4% |
|  | 1 week | 153 | 20.7% |
|  | 1 month | 197 | 26.7% |
|  | 3 months | 163 | 22.1% |
|  | 6 months | 161 | 21.8% |
|  | 1 year | 33 | 4.5% |
|  | Longer than 1 year | 22 | 3.0% |
| **Barriers to weight loss magnitude** | Diet record accuracy | 233 | 31.5% |
|  | Client nutrition proficiency | 177 | 24.0% |
|  | Diet record collection | 153 | 20.7% |
|  | Lack of weight loss results | 151 | 20.4% |
|  | Appointment scheduling | 150 | 20.3% |
|  | Low visit frequency | 138 | 18.7% |

|  |  |  |
| --- | --- | --- |
| Client retention | 130 | 17.6% |
| Others, not listed: |  |  |
| Low income | 65 | 8.8% |
| Undiagnosed eating disorders | 36 | 4.9% |
| Food insecurity | 26 | 3.5% |
| Depression | 23 | 3.1% |
| Insurance coverage for RD | 19 | 2.6% |
| Lack of adherence to prescribed diet | 16 | 2.2% |
| Reliance on fast food | 5 | 0.7% |
| Unrealistic weight loss expectations | 1 | 0.1% |

Responses to weight management outcomes and protocols questions provided by dietitians of the Weight Management Dietetic Practice Group (WMDPG) of the Academy of Nutrition and Dietetics. The total (n) number of dietitians who selected the answer and the percentage (%) of dietitians out of the total (N=739).

## 4. Discussion

This study surveyed RDNs whose primary work is dietary consultation for weight management. The survey identified the diet interventions and tools for delivering weight management, their perceived effectiveness, and their perceived barriers to client weight loss success. Key findings include a demographic characterization of weight management RDNs, a high reliance upon technology and apps by both practitioners and clients, a lack of a standard weight management program to refer to, and insufficient amount of contact with patients as barriers to successful practice.

Although surveyed RDNs reported working in various fields, they reported spending most of their time on weight management. Our surveyed RDNs in the Weight Management Dietetic Practice Group (WMDPG) as a representative population of all RDNs in the US were relatively younger, less ethnically diverse, and a higher proportion male. Surveyed RDNs were 35 years of age as opposed to the average RDN age in the US of 41 years [17]. Surveyed RDNs were also 89% white, and 14% male as opposed to the average of the US population which is 73% and 10% respectively [17].

The consequences of technology’s insertion into healthcare are notable in the results of this survey. Surveyed RDNs indicated that they primarily request diet records by smartphone application. The top three apps RDNs claimed to recommend to clients for diet and weight tracking were My Fitness Pal, Fitbit, and Calorie Count. Recent publications reflect a similar preference for the use of apps for diet recording [18, 19]. Using technology has been cited as beneficial for resolving issues with data management and interpretation, improving the ease of recording, and saving time and resources on analysis by RDNs [20]. Traditionally, multiple food records would be collected by a skilled interviewer [21]. Although our surveyed RDNs prefer using apps, they may acknowledge that relying on them is less accurate as evidenced by their identification of accurate dietary record collection as the top barrier in their weight loss practices. Concern over accuracy and efficacy of health-related apps is an area of new and growing research. New tools such as the App Quality Evaluation (AQEL) and Mobile App Rating Scale (MARS) may prove beneficial in identifying the best apps for RDNs to use with their patients [22, 23].

The majority of our surveyed RDNs claimed to not have a specific weight loss program or diet that they commonly refer to their patients, however online programs and education with generalizable information to weight loss may be warranted. For those surveyed RDNs that do rely on a specific diet or program, a low carbohydrate diet or the Obesity Medicine Program were the most prevalent. Because individual needs, taste preferences, and goals differ, it is understandable that RDNs report that they do not use a standard diet or program for their practice. However, some dietary principles for weight loss are widely applicable and could be established into a generalizable education program. For example, RDNs typically encourage an increase in protein consumption when in a calorie deficit because of its significance to preserving skeletal muscle mass [24]. Additionally, RDNs are encountering more patients who are using weight loss medications such as GLP-1 agonists that cause a rapid onset of weight loss which is more threatening to skeletal muscle mass loss. This growing number of patients may need additional diet education and enforcement outside of the counselling session [25, 26].

Most surveyed RDNs reported seeing their typical client for weight loss once a month and reported an average length of continued engagement as only one month which suggests a high rate of attrition. Elevated dropout rates in weight loss programs are an often-identified issue. Some literature hypothesizes this attrition is associated with higher weight loss expectations than actual outcomes [27–29]. Weight loss may take an initial period of experimentation prior to seeing results which can be discouraging. Another potential reason for attrition may be due to a lack of insurance coverage. While coverage for medical nutrition therapy is improving, the number of counselling sessions and length of time allotted per session varies depending on the insurer [30].

There were strengths and limitations to this survey. A strength is the number of responses, which was higher than anticipated with an estimated 3,400 total members in the WMDPG in 2022-2023 from which we recruited resulting in a total of 739 complete surveys. The survey creation and testing were rigorous including drafting from a previously published survey, the CAS, and expert content validation. A limitation to this survey is that we only provided this survey to a select group of RDNs enrolled in the WMDPG who may or may not be generalizable to those outside of this practice group. However, the objective of this study was to find practices of RDNs whose practice was primarily weight management. Additionally, the survey was provided via email and discussion board, which may bias our results towards individuals with preference for technology-assisted communication. Lastly, this survey identifies tools and interventions specific to dietary intake, but does not investigate lifestyle or behavioral interventions [31].

In conclusion, although previous research has demonstrated the significance of RDN-led weight loss coaching, this survey elucidates the barriers to current, real-world RDN practice in a quickly evolving healthcare environment. Key implications for weight management practice from the findings of this survey include an increased demand for technologically enhanced services such as telehealth, apps and online sources of information and learning. While these technologies are already in use, the accuracy of data such as dietary records remain a barrier. Additionally, contact with patients is regarded as insufficient. Further research into improved methods of data collection, reasons for patient attrition, and development of reliable education programs is warranted.

## Data Availability

The data that support the findings of this study are available from the corresponding author, AO, upon reasonable request.

## Acknowledgment

We would like to acknowledge the support of Dr. Justine Karduck who developed a survey upon which this research was role modeled as well as the dietitians who participated in the focus group studies that aided in the creation of the survey questions, Ashley Brewer, RDN, Dr. Katie Robinson, Kristina Adams, RDN, and Jennifer Burton, RDN.

## Author Contributions

Ashleigh Oliveira: conceptualization, methodology, validation, formal analysis, investigation, data curation, writing – original draft preparation, writing – review and editing, visualization, project administration Dr. Manabu T Nakamura: conceptualization, resources, writing – review and editing, supervision, funding acquisition

## Author Declarations

Author declarations: none

